# Optimal Stereotactic Body Radiotherapy Dosage For Hepatocellular Carcinoma

**DOI:** 10.1101/2020.02.27.20028621

**Authors:** Ting-Shi Su, Ying Zhou, Yong Huang, Tao Cheng, Ping Liang, Shi-Xiong Liang, Le-Qun Li

## Abstract

**Background and purpose:** The optimal dose and fractionation scheme of stereotactic body radiation therapy (SBRT) for hepatocellular carcinoma (HCC) remains unclear due to different tolerated liver volumes and degrees of cirrhosis. We compared the effectiveness of stereotactic body radiation therapy dosing regimens for hepatocellular carcinoma (HCC).

**Methods and materials:** This single-center retrospective study included 604 patients treated during 2011-2017. Biologically effective dose (BED_10_) and equivalent dose in 2 Gym fractions (EQD_2_) were assumed at an α/β ratio of 10. Overall survival (OS), local recurrence-free rate (LRF), intrahepatic recurrence-free rate (IRF), and progression-free survival (PFS) was evaluated in univariable and propensity-score matched analyses.

**Results:** Median tumor size was 5.2 cm (interquartile range [IQR], 1.1-21.0). Median follow-up was 31 months in surviving patients (IQR, 3-82). High radiotherapy dose correlated with better OS, PFS, LRF and IRF. Different post-SBRT OS, PFS, LRF and IRF rates were observed for stereotactic ablative radiotherapy (SART) with BED_10_ ≥100 Gy, SBRT with EQD_2_ ≥74 Gy to BED_10_ <100 Gy, and stereotactic conservative radiotherapy (SCRT) with EQD_2_ <74 Gy.

**Conclusions:** High radiotherapy dose correlated with better outcomes. If tolerated by normal tissue, we recommend SART as a first-line ablative dose or SBRT as a second-line radical dose. Otherwise, SCRT is recommended as palliative irradiation.

## 1. Introduction

Hepatocellular carcinoma (HCC) is highly prevalent in many Asian countries and accounts for nearly 80% of HCC cases worldwide. In China, HCC is the second most common cause of cancer-related deaths and the fourth most commonly diagnosed cancer among men [1]. HCC is resectable in only 10–40% of newly diagnosed patients. Liver resection, transplantation, percutaneous ethanol injection, or radiofrequency ablation (RFA) are the standard treatments for early-stage HCC [2].

The use of external beam radiation therapy (EBRT), specifically including stereotactic body radiation therapy (SBRT), is increasing in popularity [3-7]. It is commonly recommended as an alternative treatment in medically inoperable patients, as a result of its rapid adoption in clinical practice worldwide [8-10]. SBRT for primary HCC provides high rates of durable local control (89–100%) [4,11-14], but there is no clear evidence of a dose-survival relationship among commonly-utilized radiation therapy schedules. Increasing radiotherapy (RT) dose was associated with improved overall survival in patients treated with SBRT for stage I non-small-cell lung cancer [15-17]. However, the optimal dose and fractionation scheme of SBRT for HCC remains unclear. Therefore, in this retrospective study, we aimed to analyze the dose-survival relationship in patients undergoing SBRT for HCC by using data from a large cohort from Guangxi Zhuang Autonomous Region in China.

## 2. Results

### 2.1. Baseline characteristics

From January 1st, 2011 to March 31st, 2017, 678 primary liver cancer patients were treated with SBRT. Twenty-nine cases of intrahepatic cholangiocarcinoma, 25 cases for which complete data was lacking, and 19 cases that were lost to follow-up were excluded; finally, a total of 604 patients were enrolled in this study (Figure S1). Based on our previous studies [18,19], the RT dose was classified into three levels: stereotactic ablative radiotherapy (SART, also known as SABR) was defined as a first-line ablative dose of BED10 ≥100 Gy, SBRT was defined as a second-line radical dose of EQD2 >74 Gy to BED10 <100 Gy, and stereotactic conservative radiotherapy (SCRT) was defined as palliative irradiation of EQD2 <74 Gy. The demographic and clinical characteristics of the patients and their treatment are summarized in Table 1.

**Table 1.**
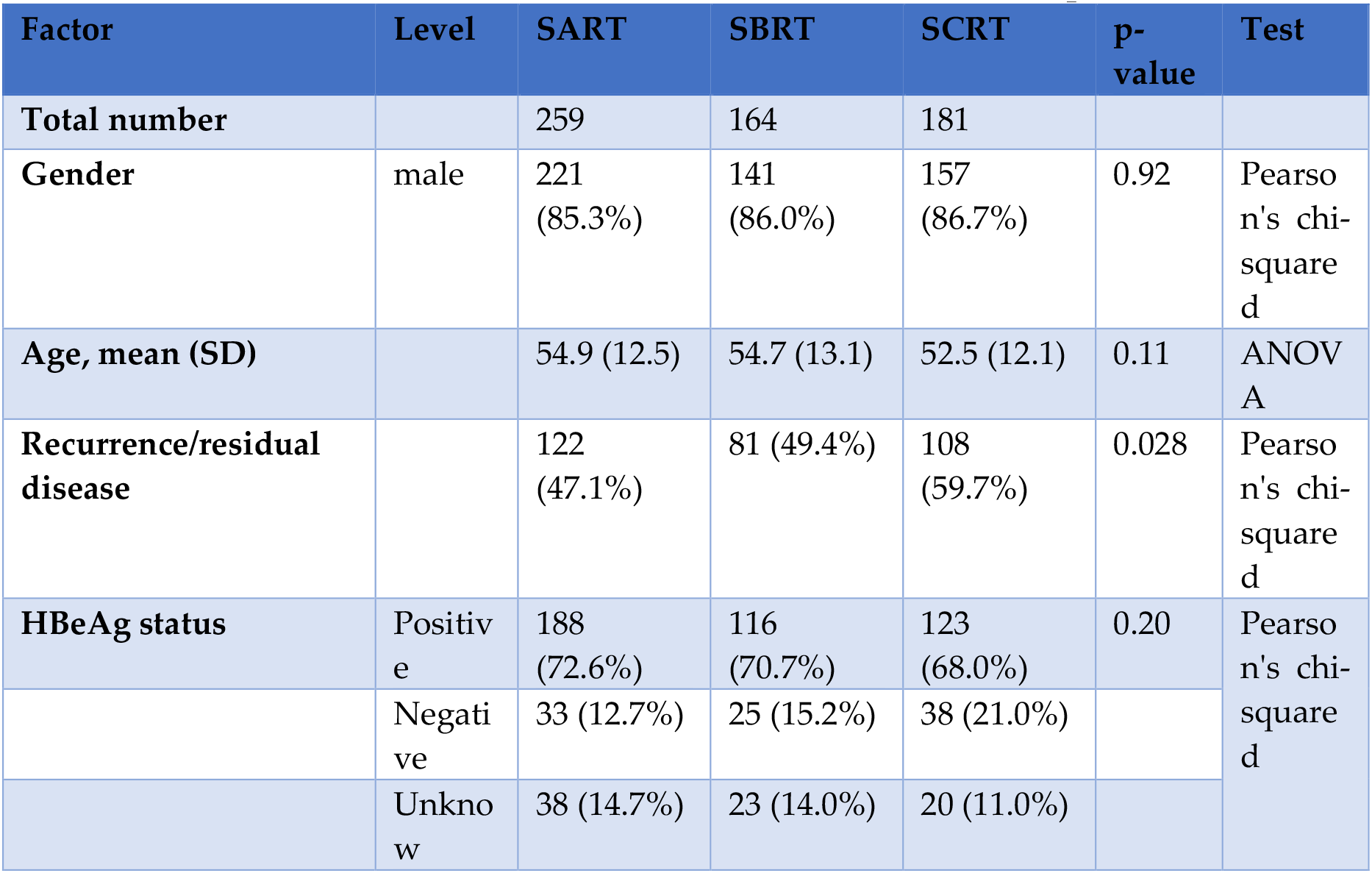
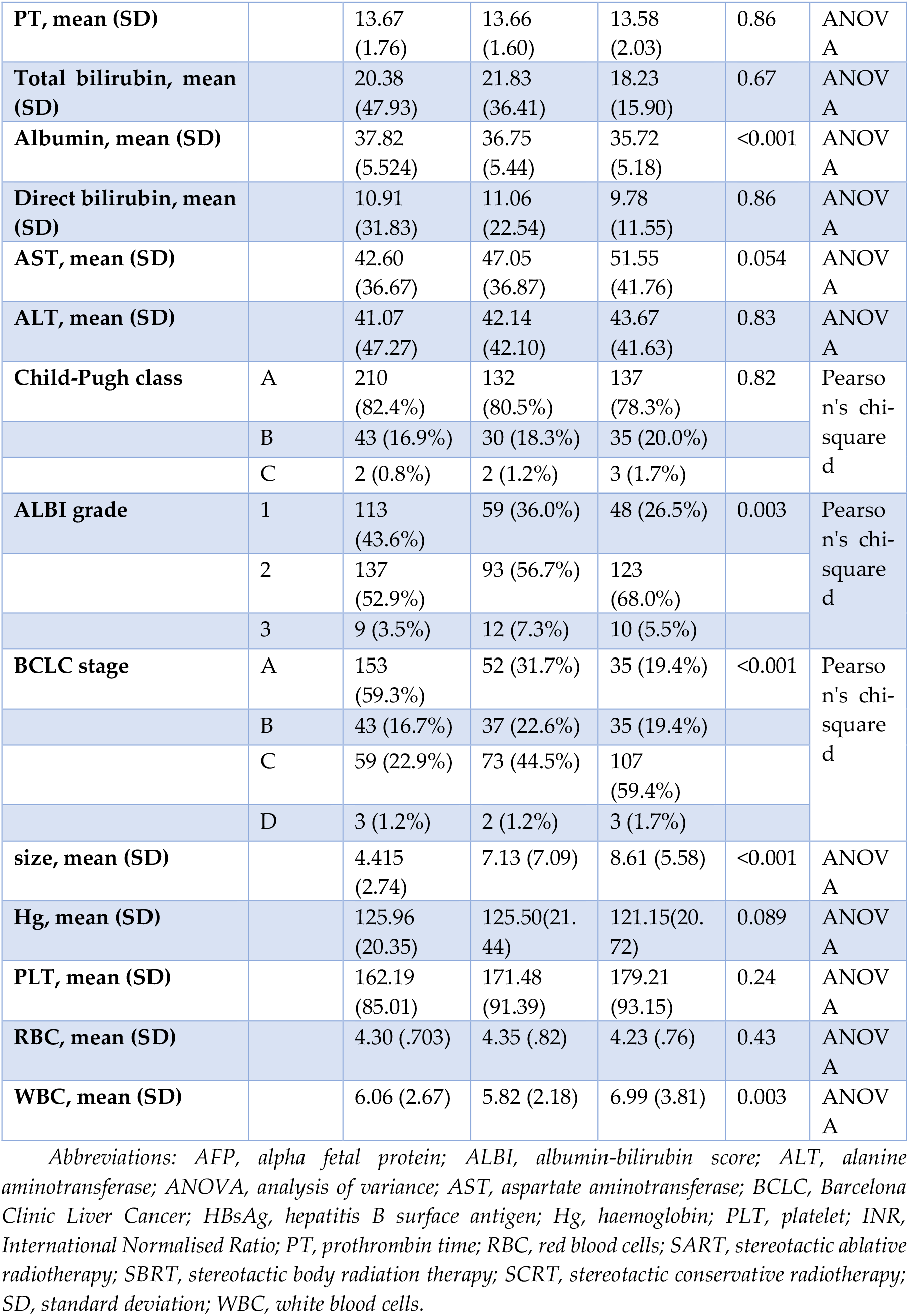
Patient and Treatment Characteristics for Different Dose Groups

### 2.2. Increasing radiation therapy dose as a prognostic factor for overall survival (OS), local recurrence-free rate (LRF), intrahepatic recurrence-free rate (IRF), and progression-free survival (PFS)

Among the included patients, the median tumor size was 5.2 cm (interquartile range [IQR], 1.1–21.0 cm). The median follow-up was 31 months in surviving patients (IQR, 3–82 months). When RT dose was used to classify the patients into the SART, SBRT, and SCRT groups, 3 notably different curves were observed for long-term post-SBRT survival. The 1-, 3-, and 5-year OS rates were 82.2%, 59.4%, and 55.3% in the SART group; 62.6%, 34.6%, and 26.4% in the SBRT group; and 49.9%, 24.5%, and 13.8% in the SCRT group, respectively (log-rank P<0.0001; Figure 1A). The 1-, 3-, and 5-year PFS rates were 58.0%, 35.1%, and 26.5% in the SART group; 40.2%, 14.3%, and 9.2% in the SBRT group; and 21.4%, 7.1%, and 5.3% in the SCRT group, respectively (log-rank P<0.0001; Figure 1B). The 1-, 3-, and 5-year LRF rates were 83.4%, 67.3%, and 63.1% in the SART group; 79.5%, 57.6%, and 57.6% in the SBRT group; and 69.5%, 54.4%, and 43.5% in the SCRT group, respectively (log-rank P=0.0141; Figure 1C). The 1-, 3-, and 5-year IRF rates were 82.2%, 59.4%, and 55.3% in the SART group; 62.6%, 34.6%, and 26.4% in the SBRT group; and 49.9%, 24.5%, and 13.8% in the SCRT group, respectively (log-rank P<0.0001; Figure 1).

**Figure 1.**
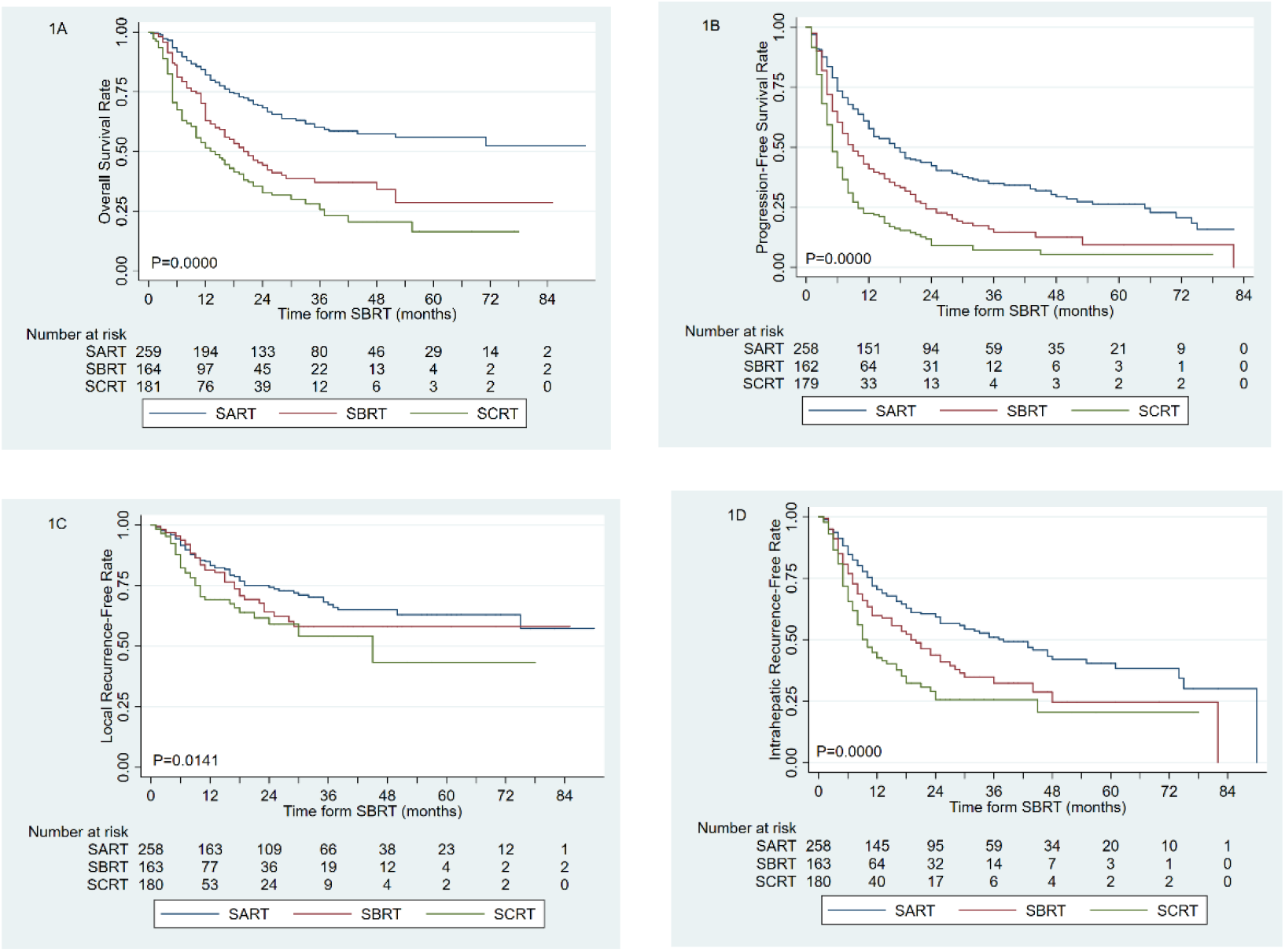
Befor propensity score matching, SART versus SBRT or SCRT: a) overall survival, b) progression-free survival, c) local recurrence-free rate, d) intrahepatic recurrence-free rate

Several variables differed substantively between the 3 groups, including the recurrence/residual disease status, level of albumin, ALBI grade, tumor size, and BCLC stage. After propensity score matching, there was no significant difference among the groups and the balance of variables was observed to be markedly improved in the matched cohorts (Tables 2 and 3).

**Table 2.** Distribution of variables in the matched cohorts SART and SBRT after propensity score matching

| Factor | Level | SART | SBRT | p-value | Test |
| --- | --- | --- | --- | --- | --- |
| N |  | 143 | 143 |  |  |
| Gender | Male | 119 (83.2%) | 123 (86.0%) | 0.51 | Pearson's chi-squared |
| Age, mean (SD) |  | 53.2 (13.0) | 54.4 (13.0) | 0.44 | Two sample t test |
| PT, mean (SD) |  | 14.0 (1.9) | 13.6 (1.6) | 0.12 | Two sample t test |
| Total bilirubin, mean (SD) |  | 23.5 (63.4) | 21.0 (37.2) | 0.68 | Two sample t test |
| Albumin, mean (SD) |  | 37.4 (5.5) | 37.0 (5.4) | 0.57 | Two sample t test |
| AST, mean (SD) |  | 45.9 (37.7) | 42.8 (29.8) | 0.44 | Two sample t test |
| ALT, mean (SD) |  | 46.4 (58.7) | 41.6 (42.2) | 0.43 | Two sample t test |
| WBC, mean (SD) |  | 6.59 (3.02) | 5.66 (2.15) | 0.022 | Two sample t test |
| RBC, mean (SD) |  | 4.22 (.68) | 4.38 (.80) | 0.18 | Two sample t test |
| PLT, mean (SD) |  | 177.6 (90.8) | 165.9 (88.9) | 0.41 | Two sample t test |
| Tumour size, mean (SD) |  | 5.30 (3.21) | 5.97 (3.10) | 0.11 | Two sample t test |
| Recurrence/residual disease | Yes | 81 (56.6%) | 72 (50.3%) | 0.29 | Pearson's chi-squared |
| HBsAg status | Positive | 94 (65.7%) | 105 (73.4%) | 0.16 | Pearson's chi-squared |
|  | Negative | 19 (13.3%) | 20 (14.0%) |  |  |
|  | Unknown | 30 (21.0%) | 18 (12.6%) |  |  |
| ALBI score, mean (SD) |  | -2.397 (.559) | -2.371 (.526) | 0.69 | Two sample t test |
| ALBI grade | 1 | 57 (39.9%) | 55 (38.5%) | 0.73 | Pearson's chi-squared |
|  | 2 | 80 (55.9%) | 79 (55.2%) |  |  |
|  | 3 | 6 (4.2%) | 9 (6.3%) |  |  |
| Total dose, mean (SD) |  | 43.6 (3.7) | 42.8 (3.1) | 0.041 | Two sample t test |
| Fraction | 1 | 6 (4.2%) | 0 (0.0%) | <0.001 | Pearson's chi-squared |
|  | 2 | 2 (1.4%) | 1 (0.7%) |  |  |
|  | 3 | 124 (86.7%) | 54 (37.8%) |  |  |
|  | 4 | 8 (5.6%) | 80 (55.9%) |  |  |
|  | 5 | 3 (2.1%) | 7 (4.9%) |  |  |
|  | 6 | 0 (0.0%) | 1 (0.7%) |  |  |
| **Child-Pugh score** | 5 | 85 (59.4%) | 77 (53.8%) | 0.75 | Pearson's chi-squared |
|  | 6 | 33 (23.1%) | 41 (28.7%) |  |  |
|  | 7 | 13 (9.1%) | 16 (11.2%) |  |  |
|  | 8 | 8 (5.6%) | 5 (3.5%) |  |  |
|  | 9 | 3 (2.1%) | 2 (1.4%) |  |  |
|  | 10 | 1 (0.7%) | 1 (0.7%) |  |  |
|  | 13 | 0 (0.0%) | 1 (0.7%) |  |  |
| **AFP status** | <8 | 41 (29.5%) | 34 (23.8%) | 0.18 | Pearson's chi-squared |
|  | 8-200 | 51 (36.7%) | 38 (26.6%) |  |  |
|  | >200 | 40 (28.8%) | 62 (43.4%) |  |  |
|  | Unknown | 7 (5.0%) | 9 (6.3%) |  |  |
| **BCLC stage** | A | 58 (40.6%) | 52 (36.4%) | 0.80 | Pearson's chi-squared |
|  | B | 35 (24.5%) | 33 (23.1%) |  |  |
|  | C | 48 (33.6%) | 56 (39.2%) |  |  |
|  | D | 2 (1.4%) | 2 (1.4%) |  |  |

**Table 3.** Distribution of variables in the matched cohorts SBRT and SCRT after propensity score matching

| Factor | Level | SBRT | SCRT | p-value | Test |
| --- | --- | --- | --- | --- | --- |
| N |  | 160 | 160 |  |  |
| **Gender** |  | 138 (86.3%) | 137 (85.6%) | 0.87 | Pearson's chi-squared |
| **Age, mean (SD)** |  | 54.5 (13.2) | 52.9 (12.1) | 0.25 | Two sample t test |
| **PT, mean (SD)** |  | 13.6 (1.6) | 13.5 (2.0) | 0.58 | Two sample t test |
| **Total bilirubin, mean (SD)** |  | 21.9 (36.8) | 17.6 (15.3) | 0.18 | Two sample t test |
| **Albumin, mean (SD)** |  | 36.7 (5.4) | 35.8 (5.1) | 0.14 | Two sample t test |
| **AST, mean (SD)** |  | 47.3 (37.2) | 52.8 (43.5) | 0.23 | Two sample t test |
| **ALT, mean (SD)** |  | 42.4 (42.5) | 43.9 (43.0) | 0.76 | Two sample t test |
| **WBC, mean (SD)** |  | 5.827 (2.18) | 7.10 (3.93) | 0.002 | Two sample t test |
| **RBC, mean (SD)** |  | 4.34 (.83) | 4.26 (.75) | 0.45 | Two sample t test |
| **PLT, mean (SD)** |  | 172.4 (92.4) | 183.6 (93.3) | 0.34 | Two sample t test |
| **Tumour size, mean (SD)** |  | 7.53 (7.15) | 8.18 (5.75) | 0.16 | Two sample t test |
| **Recurrence/residual disease** | Yes | 79 (49.4%) | 95 (59.4%) | 0.13 | Pearson's chi-squared |
| **HBsAg status** | Positive | 113 (70.6%) | 108 (67.5%) | 0.22 | Pearson's chi-squared |
|  | Negative | 24 (15.0%) | 35 (21.9%) |  |  |
|  | Unknown | 23 (14.4%) | 17 (10.6%) |  |  |
| **ALBI score, mean (SD)** |  | -2.338 (.534) | -2.281 (.507) | 0.33 | Two sample t test |
| **ALBI grade** | 1 | 57 (35.6%) | 43 (26.9%) | 0.11 | Pearson's chi-squared |
|  | 2 | 91 (56.9%) | 110 (68.8%) |  |  |
|  | 3 | 12 (7.5%) | 7 (4.4%) |  |  |
| **Total dose, mean (SD)** |  | 42.5 (3.1) | 40.1 (4.6) | <0.001 | Two sample t test |
| **Fraction** | 2 | 1 (0.6%) | 0 (0.0%) | <0.001 | Pearson's chi-squared |
|  | 3 | 68 (42.5%) | 39 (24.4%) |  |  |
|  | 4 | 83 (51.9%) | 75 (46.9%) |  |  |
|  | 5 | 7 (4.4%) | 42 (26.3%) |  |  |
|  | 6 | 1 (0.6%) | 4 (2.5%) |  |  |
| **Child-Pugh score** | 5 | 83 (51.9%) | 75 (46.9%) | 0.52 | Pearson's chi-squared |
|  | 6 | 46 (28.7%) | 52 (32.5%) |  |  |
|  | 7 | 18 (11.3%) | 21 (13.1%) |  |  |
|  | 8 | 7 (4.4%) | 7 (4.4%) |  |  |
|  | 9 | 4 (2.5%) | 1 (0.6%) |  |  |
|  | 10 | 1 (0.6%) | 2 (1.3%) |  |  |
|  | 11 | 0 (0.0%) | 2 (1.3%) |  |  |
|  | 13 | 1 (0.6%) | 0 (0.0%) |  |  |
| **AFP status** | <8 | 37 (23.1%) | 35 (21.9%) | 0.90 | Pearson's chi-squared |
|  | 8-200 | 39 (24.4%) | 37 (23.1%) |  |  |
|  | >200 | 75 (46.9%) | 81 (50.6%) |  |  |
|  | Unknown | 9 (5.6%) | 7 (4.4%) |  |  |
| **BCLC stage** | A | 48 (30.0%) | 34 (21.3%) | 0.20 | Pearson's chi-squared |
|  | B | 37 (23.1%) | 33 (20.6%) |  |  |
|  | C | 73 (45.6%) | 90 (56.3%) |  |  |
|  | D | 2 (1.3%) | 3 (1.9%) |  |  |

For the comparison of SART and SBRT, the matching procedure resulted in the selection of 143 patient pairs. The 1-, 3-, and 5-year OS rates were 76.3%, 50.6%, and 48.0% in the SART group and 65.8%, 36.3%, and 33.0% in the SBRT group, respectively (log-rank P=0.0167; Figure 2A). The 1-, 3-, and 5-year PFS rates were 51.7%, 31.4%, and 25.6% in the SART group and 43.2%, 15.3%, and 9.9% in the SBRT group, respectively (log-rank P=0.0058; Figure 2B). The 1-, 3-, and 5-year LRF rates were 85.8%, 69.5%, and 64.0% in the SART group and 80.5%, 63.8%, and 57.6% in the SBRT group, respectively (log-rank P=0.2008; Figure 2C). The 1-, 3-, and 5-year IRF rates were 68.4%, 51.4%, and 43.8% in the SART group and 59.0%, 30.4%, and 26.6% in the SBRT group, respectively (log-rank P=0.0051; Figure 2D)

**Figure 2.**
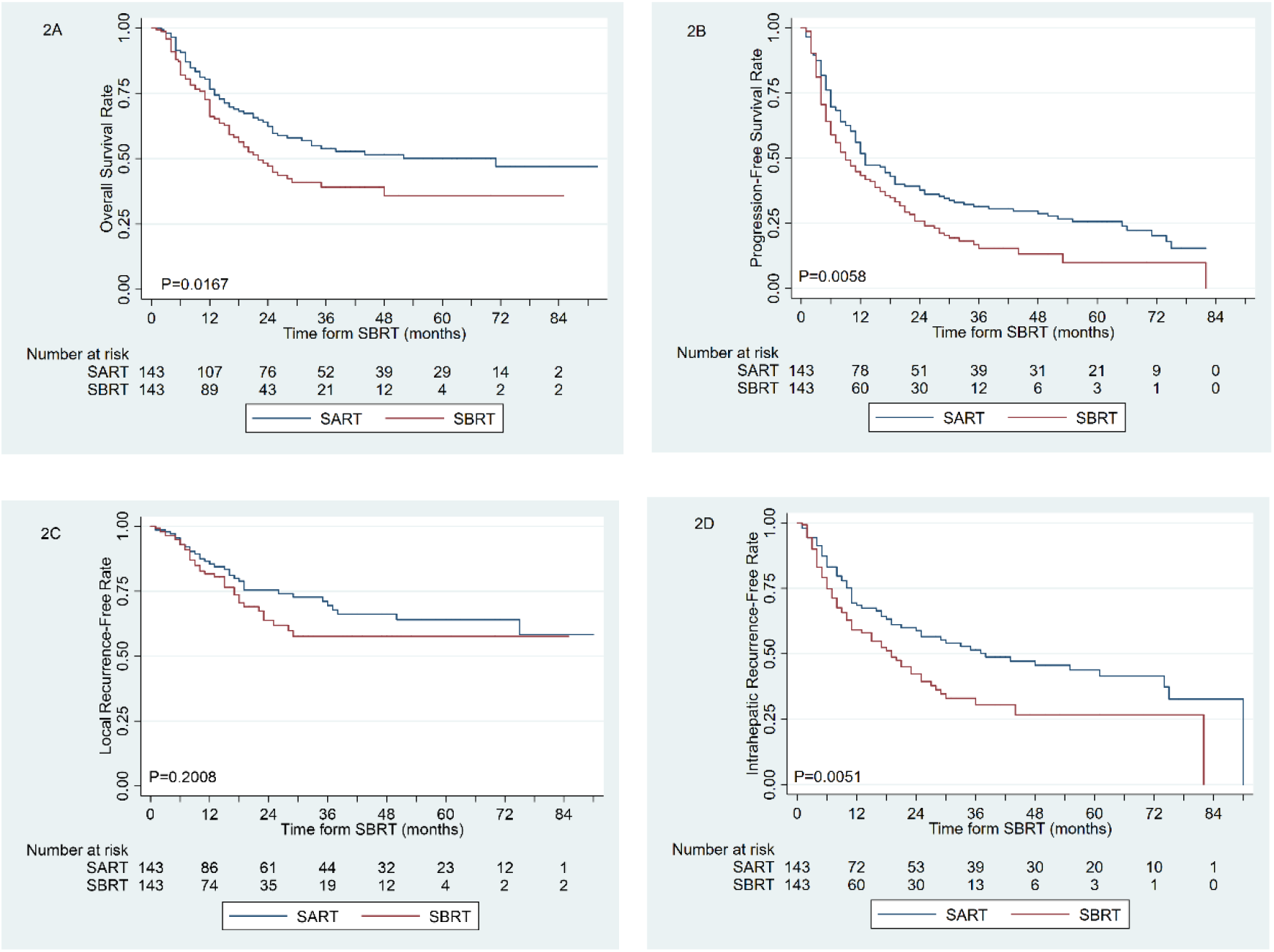
After propensity score matching, SART versus SBRT: a) overall survival, b) progression-free survival, c) local recurrence-free rate, d) intrahepatic recurrence-free rate.

For the comparison of SBRT and SCRT, the matching procedure resulted in the selection of 160 patient pairs. The 1-, 3-, and 5-year OS rates were 71.6%, 33.7%, and 25.7% in the SBRT group and 49.9%, 24.8%, and 13.9% in the SCRT group, respectively (log-rank P=0.0158; Figure 3A). The 1-, 3-, and 5-year PFS rates were 41.3%, 13.7%, and 8.8% in the SBRT group and 22.3%, 7.0%, and 5.3% in the SCRT group, respectively (log-rank P=0.0004; Figure 3B). The 1-, 3-, and 5-year LRF rates were 80.9%, 57.2%, and 57.2% in the SBRT group and 70.7%, 55.9%, and 44.7% in the SCRT group, respectively (log-rank P=0.116; Figure 3C). The 1-, 3-, and 5-year IRF rates were 58.8%, 31.3%, and 23.8% in the SBRT group and 44.9%, 26.3%, and 21.0% in the SCRT group, respectively (log-rank P=0.041; Figure 3D).

**Figure 3.**
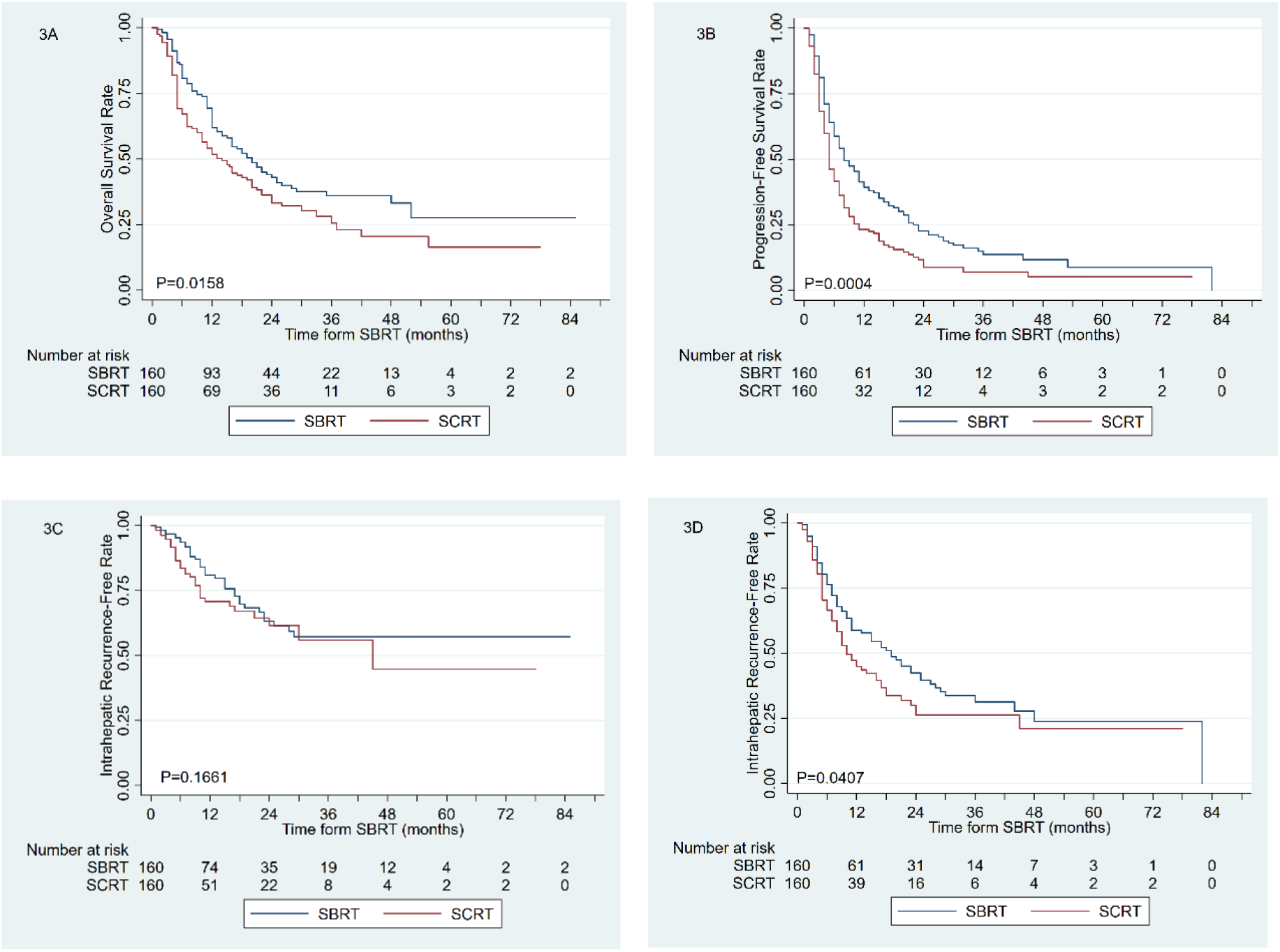
After propensity score matching, SBRT versus SCRT: a) overall survival, b) progression-free survival, c) local recurrence-free rate, d) intrahepatic recurrence-free rate.

### 2.3. Subgroup analysis of radiation therapy total dose for OS

Additionally, we found a significant association between higher total dose (TD) and better OS. The 1-, 3-, and 5-year OS rates were 71.4%, 49.7%, and 43.6% in the TD ≥42 Gy group and 56.5%, 23.5%, and 20.5% in the TD <42 Gy group, respectively (log-rank P<0.0001; Figure 4A). Further, we found that the use of fewer fractions was association with significantly better OS. The 1-, 3-, and 5-year OS rates were 74.6%, 47.8%, and 45.3% in the ≤3 fractions group and 56.7%, 35.1%, and 20.3% in the ≥4 fractions group, respectively (log-rank P=0.0002; Figure 4B).

**Figure 4.**
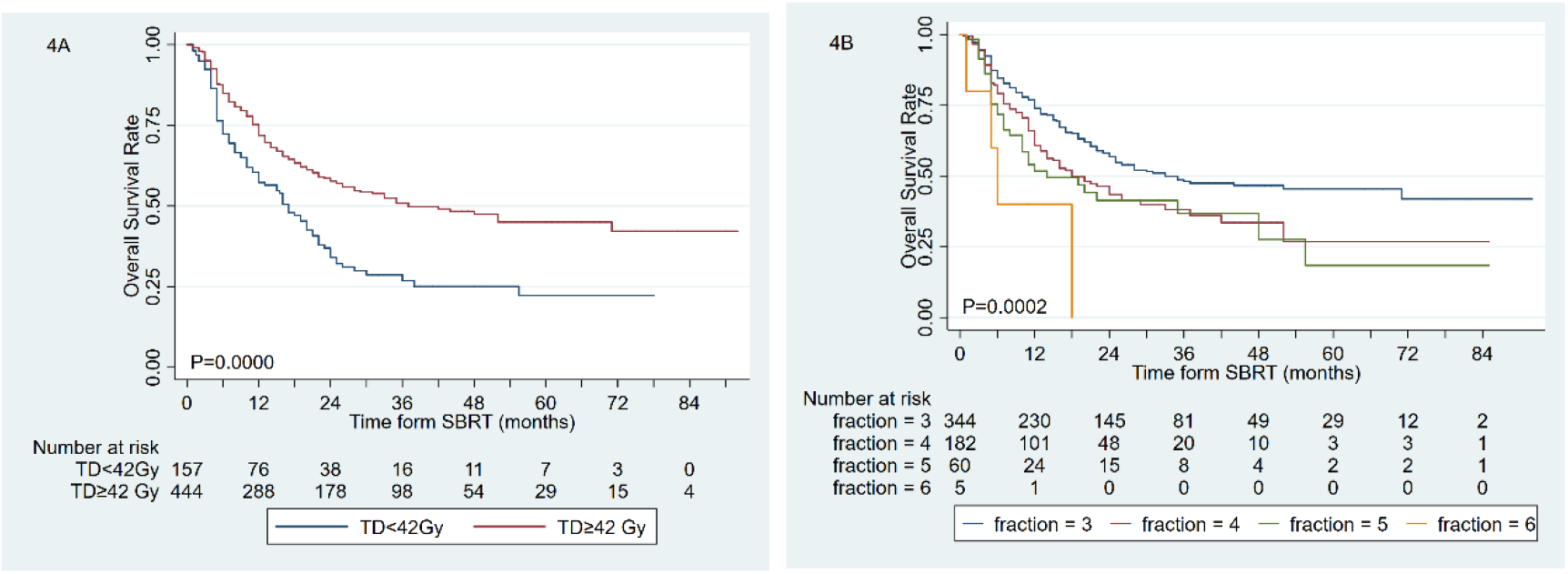
Different post-SBRT overall survival: a), Total doses ≥42 Gy versus <42 Gy group; b) fractions ≤3 versus ≥4 fractions group.

### 2.4. Multivariable analysis of radiation therapy dose for OS

In a multivariable analysis, high RT dose was associated with significantly improved OS (P<0.05). The other clinical factors that had significant associations with OS were age ≥60 years, ALBI scores, alanine aminotransferase (ALT), and BCLC stage (all P<0.05, Table 4).

**Table 4.** Multivariable predictors of overall survival

| Factor | level | HR | SE | P | [95% Conf. Interval] |  |
| --- | --- | --- | --- | --- | --- | --- |
| Gender | male | 1.000 |  |  |  |  |
|  | female | 0.843 | 0.173 | 0.405 | 0.565 | 1.259 |
| Age | $< 60$ | 1.000 | | | | |
| | $\geq 60$ | 1.424 | 0.241 | 0.036 | 1.023 | 1.984 |
| BCLC stage | A | 1.000 |  |  |  |  |
|  | B | 2.196 | 0.518 | 0.001 | 1.384 | 3.486 |
|  | C | 3.555 | 0.724 | 0.000 | 2.385 | 5.300 |
|  | D | 1.929 | 1.259 | 0.314 | 0.537 | 6.929 |
| HCC status | recurrence/residual | 1.000 |  |  |  |  |
|  | primary | 0.850 | 0.131 | 0.290 | 0.629 | 1.149 |
| AFP status | $< 8$ | 1.000 | | | | |
|  | 8-200 | 0.662 | 0.406 | 0.501 | 0.199 | 2.201 |
| | $> 200$ | 0.944 | 0.574 | 0.924 | 0.286 | 3.109 |
|  | Unknown | 1.213 | 0.728 | 0.747 | 0.374 | 3.933 |
| RT dosage | SART | 1.000 |  |  |  |  |
|  | SBRT | 1.506 | 0.312 | 0.048 | 1.003 | 2.260 |
|  | SCRT | 1.846 | 0.397 | 0.004 | 1.212 | 2.813 |
| ALBI scores |  | 1.760 | 0.340 | 0.003 | 1.206 | 2.569 |
| Size |  | 1.004 | 0.025 | 0.861 | 0.957 | 1.054 |
| AST |  | 1.003 | 0.002 | 0.177 | 0.999 | 1.006 |
| ALT |  | 0.995 | 0.002 | 0.022 | 0.990 | 0.999 |
| WBC |  | 1.041 | 0.025 | 0.088 | 0.994 | 1.091 |
| RBC |  | 1.049 | 0.138 | 0.714 | 0.810 | 1.359 |
| PLT |  | 1.000 | 0.001 | 0.883 | 0.998 | 1.002 |
| Hg |  | 0.992 | 0.005 | 0.126 | 0.982 | 1.002 |

## Discussion

Precise radiotherapy dose is important but uncertain, especially in HCC that can be treated with radical radiotherapy, because primary HCCs tend to be associated with different tolerated liver volumes and degrees of cirrhosis. In the current study, we classified RT doses into three levels (SART, SBRT, and SCRT) and found that higher RT doses were associated with better OS in both univariable and multivariable analyses. After propensity score matching, BED10 ≥100 Gy and EQD2 ≥74 Gy continued to be associated with significantly better OS and PFS, which suggests that they conferred significant survival benefits. This finding is consistent with a previous study of SBRT for HCCs that were >5 cm and lacked tumor thrombus, lymph node involvement, and extrahepatic metastasis. In that study, BED10 ≥100 Gy and EQD2 ≥74 Gy were significant prognostic factors for OS, progression-free survival, local relapse-free survival, and distant metastasis-free survival [18]. Scorsetti et al. [20] studied 43 patients with HCC (1–3 lesions, ≤6 cm) who received SBRT and found that higher BED10 was significantly correlated with improved local control and OS. The 1-year local control rate was 100% in patients treated with BED10 ≥100 Gy, while it was 52.0% in patients treated with BED10 <100 Gy, with median OS times of 27.0 versus 8.1 months (P<0.05). We have speculated that individualized doses of SBRT might be effective for HCC because primary HCCs tend to be associated with different degrees of cirrhosis and liver volumes. In our study, RT doses were classified into three levels: SART, SBRT and SCRT. If tolerated by normal tissue, a first-line ablative dose of SART with a BED10 ≥100 Gy or a second-line radical dose of SBRT with an EQD2 ≥74 Gy is recommended. Otherwise, palliative irradiation via SCRT with EQD2 <74 Gy is recommended.

Wahl et al. [4] reported that SBRT appears to be a reasonable first-line treatment for inoperable large HCC. They found no significant difference in OS between the SBRT and RFA groups. One-year OS was 74% after SBRT and 70% after RFA, while 2-year OS was 46% after SBRT and 53% after RFA. Wahl et al. also observed that, for tumors sized ≥2 cm, SBRT was superior to RFA in terms of freedom from local progression. On the other hand, Rajyaguru et al. [21] reported unfavorable OS in patients receiving SBRT, with 5-year OS rates of 19.3% in the SBRT group and 29.8% in the RFA group. Their study included a relatively small sample population of patients receiving treatment with SBRT (n=275), and the data for their study was retrieved from a US national cancer database. RT dose was unknown for 40 (14%) of the 275 patients receiving SBRT. Of the 235 with known doses, 60 (26%) received lower radiation doses (<40 Gy). A follow-up analysis of patients receiving ablative doses showed no OS difference in comparison with patients receiving RFA [22]. In the current study, which had a larger sample population (n=604), the 1-, 3-, and 5-year OS rates were 82.2%, 59.4%, and 55.3% in the SART group; 62.6%, 34.6%, and 26.4% in the SBRT group; and 49.9%, 24.5%, and 13.8% in the SCRT group, respectively (P<0.0001). In a previous study of the same patient cohort, there was no difference in long-term survival outcomes between SABR and liver resection for small HCCs (≤5 cm) with CP-A cirrhosis. The 5-year OS rates were 70.0% and 64.4% in the in the SABR and liver resection groups, respectively [5]. Therefore, precise radiotherapy dose is important, especially in HCC that can be treated with radical radiotherapy.

In the current study, a ≥42-Gy total dose and ≤3 fractions were also important indices that were associated with clinical curative effect. This result may be beneficial to the clinical application of SBRT in the treatment of HCC, considering that this has been rapidly adopted as a preferred treatment choice worldwide. Jang et al. [23] reported SBRT doses escalated from 33 Gy in 3 fractions to 60 Gy in 3 fractions for HCC (longest diameter ≤7 cm). The 2-year OS rates for patients treated with doses >54 Gy, 45–54 Gy, and <45 Gy were 71%, 64%, and 30%, respectively, while the 2-year local control rates were 100%, 78%, and 64%, respectively. Six patients showed a deterioration in the CP score to ≥2 within 3 months, and 5 patients experienced grade ≥3 gastrointestinal toxicity. In our clinical practice in China, a fractionated scheme of 45 Gy in 3 fractions (BED10=112.5 Gy and EQD2=93.8 Gy) was applied in a multi-institutional, single-arm Phase II trial of SBRT for the treatment of HCC in patients with unifocal liver tumors within ≤5 cm in diameter, or 2–3 lesions with a maximum lesion size of 3 cm for each lesion (NCT 02363218). Thirteen patients presented with grade ≥2 hepatic adverse reaction and 8 patients presented with decreases in CP classification. Total liver volume >1,162 mL and normal liver volume >1,148 mL should be ensured to improve therapeutic safety [24]. Additionally, another fractionated scheme of 39–50 Gy in 3–5 fractions were applied in our single-institutional Phase II trial of SBRT for the treatment of HCC in patients with 1–3 nodular HCC lesions with a total diameter <10 cm (ChiCTR-IIC-16008233). The dose-volume constraints for the liver were the absolute normal liver volume spared from at least 15 Gy (VS15) >700 mL and/or the percentage (%) of normal liver volume receiving more than 15 Gy (V15) <1/3 normal liver volume. If tolerated by normal tissue, a first-line ablative dose of SART with a BED10 ≥100 Gy or a second-line radical dose of SBRT with an EQD2 ≥74 Gy was recommended. Otherwise, palliative irradiation via SCRT with EQD2 <74 Gy was recommended. No case of classic radiation-induced liver disease was observed. Regarding the Child-Pugh (CP) scores following SBRT, 20 (23.5%) and 12 (14.2%) patients had CP+ ≥1 and ≥2, respectively. We further found that V15, VS10 (the absolute normal liver volume spared from at least 10 Gy), and pre-CP were optimal predictors for radiation-induced hepatic toxicity (RIHT: CP+ ≥1 and ≥2) modelling. We built normal tissue complication probability models and nomograms for RIHT predication in order to obtain individual constraints for each patient. VS10 ≥416.2 mL and V15 <33.1% for RIHT (≥1) risk stratification, and VS10 ≥621.8 mL and V15 < 21.5% for RIHT (≥2) may provide some references for liver constraint selection (table 5) [19].

**Table 5.** Recommendations for 3-5 fractions SBRT treatment

| Dosimetric constraints for liver | Radiation dose for GTV |
| --- | --- |
| V15 $<21.5\%$, VS10 $\geq 621.8$ mL | SART: BED10 $\geq 100$ Gy |
| V15 $<33.1\%$, VS10 $\geq 416.2$–$621.8$ mL | SBRT: EQD2 $\geq 74$ Gy |
| Without above conditions or Child-Pugh $\geq B7$ class | SCRT: EQD2 $<74$ Gy |

BED10 can serve as a simple and straightforward means to perform a comparative and effective analysis among a large variety of dose fractionations prescribed. The clinical efficacy of higher BED10 values has been fully recognized in the use of SBRT for the treatment of lung cancer. Kestin et al. [16] analyzed 505 patients receiving SBRT for the treatment of non-small-cell lung cancer at 5 institutions, and found that local recurrence was significantly decreased for those treated with BED10 <105 Gy as compared to those treated with BED10 >105 Gy (15% vs. 4%). Onishi et al. [17] analyzed 257 patients at 14 institutions treated with SBRT for lung cancer and found a 5-year local control of 37% versus 84% for patients treated with an isocenter of BED10 <100 Gy versus BED10 >100 Gy. Koshy et al. [15] analyzed 498 patients receiving SBRT for early stage non-small-cell lung cancer, and found a survival benefit with BED10 >150 Gy in T2 tumor patients.

The present study has some limitations. First, the calculation of BED10 using an α/β ratio of 10 from the linear-quadratic model is controversial, even though it is commonly used. Second, our study had a single-center retrospective design, and used data from a larger cohort of patients from Guangxi Zhuang Autonomous Region in China. Third, we observed strong associations between RT dose/fractionation and other prognostic factors, including BCLC class, tumor size, and ALBI grade. We have speculated that individualized doses of SBRT might be effective for HCC because primary HCCs tend to be associated with different degrees of cirrhosis and liver volumes. In general, patients with small tumors, BCLC stage A, and/or low ALBI score received higher RT doses, whereas those with larger tumors, BCLC B, C, D, and/or higher ALBI score received lower RT doses. This selection bias may not have been completely addressed with the multivariable analysis. For this reason, we also applied propensity score matching as an additional means of reducing selection bias and confounding. Applying propensity score matching created a quasi-randomized experiment. After matching, there was no significant difference in pre-RT covariates among the treatment groups and the balance of variables in the matched cohorts was markedly improved, but the associations between higher doses and better OS and PFS remained, suggesting that they are not the product of biases. Fourth, Short-term liver toxicity has been reported, but classic radiation-induced liver disease was not observed during the follow-up period. SBRT dosing regimens for HCC are generally tolerable, and further research is being conducted on long-term OS and liver toxicity. ChiCTR-IIC-16008233 (39–50 Gy in 3–5 fractions based on SART, SBRT, SCRT) and NCT 02363218 (45Gy in 3 fractions) are ongoing clinical trials in our hospital in China. Fifth, this study was performed in an area in which hepatitis B is endemic; therefore, it is unclear whether the dosimetric findings are applicable to HCC cases associated with other risk factors.

## 4. Materials and Methods

### 4.1. Patients

A dataset collected from Guangxi Zhuang Autonomous Region in China was used in this retrospective study. Radiation therapy for liver cancer has been used in our region for more than 20 years [5,25]. HCC diagnosis was established based on histopathology or according to the clinical criteria for diagnosis of HCC [9]. The eligibility criteria were as follows: patients with primary HCC treated with SBRT from January 1, 2011 to March 31, 2017. The exclusion criteria were as follows: (a) prior history of abdominal conventional radiotherapy, (b) intrahepatic cholangiocellular carcinoma, (c) gallbladder metastases, and/or (d) liver metastases. This study complied with the tenets of the Declaration of Helsinki. All study participants provided informed consent, and the study design was approved by the appropriate ethics review board.

### 4.2. Stereotactic body radiation therapy

The SBRT technique used at our institution has been previously described [5,12,18,19]. Briefly, the patients were immobilized with a customized external vacuum-type. SBRT was delivered using the CyberKnife system (Accuray Incorporated, Sunnyvale, CA, USA), with 6 Mv photons. Three or four gold markers were inserted into the surrounding area of the tumor or into tumor tissue. Gross tumor volume (GTV) was delineated as the visible tumor, based on results of computed tomography and/or magnetic resonance imaging. Planning target volume (PTV) was established as a 0–5 mm expansion of the GTV. No internal target volume (ITV) was created because tracking was used. A dose of 28–55 Gy was administered in 1–6 fractions on consecutive days at the 50–85% isodose line that covered at least 97% of the PTV. Total doses and fractionation schedules were chosen according to size and dose-volume constraints of the organs at risk.

### 4.3. Calculated values

Biologically effective dose (BED10) and equivalent dose in 2 Gy fractions (EQD2) were assumed at an α/β ratio of 10, for rapidly proliferating tumor cells. EQD was calculated as: d×n {(α/β+d)/(α/β+dx)}; BED was calculated as: d×n{1+d/(α/β)}; (d=dose, n=fraction and dx=2). OS rates were calculated from the date of initiation of SBRT until the date of final follow-up or death.

### 4.4. Statistical analysis

OS, PFS LRF, and LFR rates were estimated using the Kaplan–Meier method and compared between groups using the log-rank test. OS was calculated starting from the date of the first treatment until the date of the final follow-up or death. LFR was calculated starting from the date of the first treatment until the date of local recurrence or progression. IFR was calculated starting from the date of the first treatment until the date of intrahepatic recurrence or progression. PFS was calculated starting from the date of the first treatment until the date of recurrence or progression or death. Additionally, multivariate regression analyses of survival were performed using the Cox proportional hazards model. The multivariate regression included all characteristics that were significant in univariate analyses, as well as clinical characteristics that could be associated with survival based on previously published evidence. Student’s t and Mann-Whitney U tests were used to analyze continuous variables. For categorical variables, the χ2 and Fisher’s exact tests were used. In our analyses of the effects of treatments, propensity score matching methods were applied to reduce the potential for selection bias and confounding. Matching was performed on a 1:1 basis and the caliper value for matching was set to 0.1. Age, sex, tumor size, albumin-bilirubin (ALBI) score, and Barcelona Clinic Liver Cancer (BCLC) stage were selected on the basis of this score, and the values were compared with those observed at the baseline. Statistical analyses were performed using Stata 15.1 (StataCorp LLC, USA) and SPSS 23.0 (IBM, Armonk, NY). A P-value less than 0.05 was considered statistically significant.

## 5. Conclusions

In this large retrospective study, higher RT doses were associated with better survival in patients undergoing SBRT for the treatment of HCC. Individualized doses of SBRT are recommended because patients with HCC have different degrees of cirrhosis and liver volumes. If tolerated by normal tissue, we recommend SART with BED10 ≥100 Gy as the first-line ablative dose or SBRT with EQD2 ≥74 Gy and BED10 <100 Gy as the second-line radical dose. Otherwise, SCRT with EQD2 <74 Gy is recommended as palliative irradiation. Further research is warranted to validate the effects of this novel treatment regimen, and would preferably include multi-institutional prospective studies.

## Data Availability

None

## Author Contributions

Su TS made substantial contributions to conception and design of the study. Su TS, Liang P, Zhou Y, Cheng T and Huang Y made substantial contributions to data acquisition. Su TS made substantial contributions to the analysis and interpretation of the data. All authors participated in drafting the article or revising it critically for important intellectual content. All authors provided final approval of the version to be published.

## Acknowledgements

We thank all patients who participated in this study and our colleagues in Rui Kang Hospital, such as Zuping Lian, Qiuhua Liu and Encun Hou and so on.

